# The estimated disease burden of COVID-19 in Japan from 2020 to 2021

**DOI:** 10.1101/2022.12.14.22283492

**Authors:** Shinya Tsuzuki, Philippe Beutels

## Abstract

**Background:** To date, it is not fully understood to what extent COVID-19 has burdened society in Japan. This study aimed to estimate the total disease burden due to COVID-19 in Japan during 2020-2021.

**Methods:** We stratify disease burden estimates by age group and present it as absolute Quality Adjusted Life Years (QALYs) lost and QALYs lost per 100,000 persons. The total estimated value of QALYs lost consists of (1) QALYs lost brought by deaths due to COVID-19, (2) QALYs lost brought by inpatient cases, (3) QALYs lost brought by outpatient cases, and (4) QALYs lost brought by long-COVID.

**Findings:** QALYs lost due to COVID-19 was estimated as 286,781·7 for two years, 114·0 QALYs per 100,000 population per year. 71·3% of them were explained by the burden derived from deaths. Probabilistic sensitivity analysis showed that the burden of outpatient cases was the most sensitive factor.

**Interpretation:** The large part of disease burden due to COVID-19 in Japan from the beginning of 2020 to the end of 2021 was derived from Wave 3, 4, and 5 and the proportion of QALYs lost due to morbidity in the total burden increased gradually. The estimated disease burden was smaller than that in other high-income countries. It will be our future challenge to take other indirect factors into consideration.

**Fundings:** This research was funded by JSPS KAKENHI [Grant number 20K10546]. The funders had no role in study design, data collection and analysis, decision to publish, or preparation of the manuscript.

## Introduction

Coronavirus disease 2019 (COVID-19) caused by the SARS-CoV-2 virus, has become a global health threat since the beginning of 2020^1–3^. In Japan, it was first detected in early 2020^4^.

This emerging infectious disease became one of the most pressing concerns for the Japanese general population and the Ministry of Health, Labour and Welfare (MHLW) Japan in early 2020 and the Prime Minister of Japan declared the state of emergency on 7^th^ April 2020 for seven prefectures including the Tokyo metropolitan area^5–7^. MHLW recommended to avoid Three Cs (Closed spaces, Crowded places, and Close-contact settings) to prevent COVID-19 transmission^8^ and behaviour of the general population drastically changed. The number of healthcare facility visits and the consumption of antimicrobials decreased substantially after the emergence of COVID-19 in Japan^9,10^. In short, the COVID-19 pandemic changed the Japanese way of life substantially.

To date, it is not fully understood to what extent this novel emerging disease has burdened society. A quantification of the observed burden, despite the great efforts that were made to minimise it, is a first step towards understanding how pandemic management can be improved.

Compared with other high-income countries, the cumulative incidence of COVID-19 cases, hospitalisations and deaths has been comparatively small in Japan, at least until the end of the year 2021^11^. For instance, the United Kingdom reported 952·6 cumulative hospitalizations per 100,000 population^12^ and the United States reported 11,700·6 cumulative hospitalizations per 100,000 population at the end of 2021^13^, while Japan reported 1,706·1 cumulative hospitalizations per 100,000 population in the same period. As for deaths, Japan reported lower rates (14·6 deaths per 100,000 population) compared with the UK and the US (218·1 and 247·8 deaths per 100,000 population, respectively) and the average of the world (69·1 deaths per 100,000 population)^14^.

In order to learn from the crisis and be better prepared for future pandemics, we aim to assess the burden caused by COVID-19 in more detail. We can classify the disease burden directly caused by COVID-19 into four categories;

i. Quality Adjusted Life Year (QALY) losses caused by fatal cases
ii. QALY losses caused by outpatient cases
iii. QALY losses caused by mild to severe inpatient cases
iv. QALY losses caused by long-COVID^15^

Since they can be both interpreted as indicators of pandemic management, here we aim to document both the cumulative and the chronological, per-wave, disease burden caused by COVID-19. The COVID-19 epidemic in Japan was characterised by five waves in 2020 and 2021. We adopted a previously proposed classification of waves observed in Japan^16^, as follows (see also Figure 2):

i. First wave (Wave 1), 01/01/2020-05/31/2020;
ii. Second wave (Wave 2), 06/01/2020-10/31/2020;
iii. Third wave (Wave 3), 11/01/2020-03/31/2021;
iv. Fourth wave (Wave 4), 4/1/2021-6/30/2021;
v. Fifth wave (Wave 5), 7/1/2021-12/31/2021

The Japanese government had implemented different non-pharmaceutical interventions (NPIs) in each period and the guidelines for clinical management of COVID-19 cases grew gradually, implying the characteristics of the burden in each wave are expected to be different.

The main objective of this study is to assess the disease burden caused by COVID-19 in Japan between the beginning of 2020 and the end of 2021 in order to enable comparisons over time, with other diseases and with other countries.

## Methods

### Settings

We constructed a progression pathway model of COVID-19 infection (Figure 1), in which two types of infection; symptomatic and asymptomatic, and three degrees of severity were defined; outpatient cases, inpatient cases (mild), and inpatient cases (severe). The definition of “severe” inpatient cases varied by prefecture because each prefecture defined the severity of inpatient COVID-19 cases by its own criteria. A large part of prefectures defined “severe” cases as patients admitted to an intensive care unit (ICU) or patients requiring mechanical ventilation. Additionally, we assumed that any type of symptomatic infection could lead to long-COVID. In line with previous studies^17,18^, we defined long-COVID patients as presenting with COVID-19 symptoms for longer than four weeks from symptom onset. The final stage of each infected case was “Death” or “Recovery”. We assumed that all symptomatic infections (both outpatient and inpatient cases), including long-COVID episodes, and deaths contributed to the disease burden, whereas asymptomatic infections did not.

**Figure 1.**
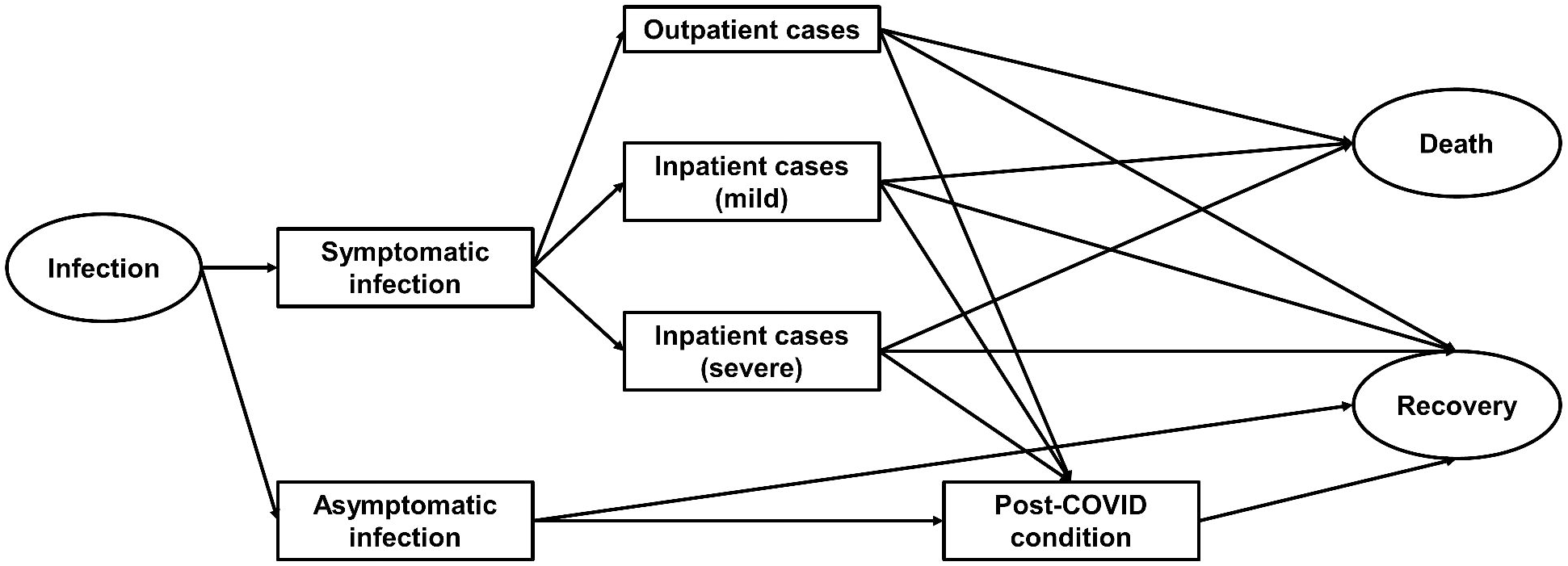
Progression pathway diagram of COVID-19 infection.

We estimated the disease burden due to COVID-19 in Japan for the period from the beginning of 2020 to the end of 2021 because the first case of COVID-19 in Japan was detected on 15^th^ January 2020^4^, and by the end of 2021 80·4% of the population had received their primary course of COVID-19 vaccination. Additionally, the less severe Omicron variant of concern (VOC), became dominant early in 2022, and its clinical, detection and epidemiological characteristics were quite different from other strains.

### Data sources

We used open data sources for the daily number of confirmed cases and deaths^19^. Demographic data were sourced from official statistics^20^. Disutility of each health status was presented as QALYs lost per episode and defined according to values from the literature^17,21,22^. We assumed the proportion of acute symptomatic COVID-19 cases that gives rise to long-COVID was 16·6% in adults and 3·9% in children^23,24^. An overview of these parameters is shown in Table 1.

**Table 1.** Details of parameters included in the model.

| Parameters | Value | Distribution | Reference |
| --- | --- | --- | --- |
| **Disutility (QALYs lost)** |  |  |  |
| **Outpatient case** | 0.033 | Lognormal<br>(mean = -5.187, SD = 0.034) | 21 |
| **Inpatient case** | 0.439 | Normal<br>(mean = 0.439, SD = 0.027) | 22 |
| **Long-COVID** | 0.129 | Lognormal<br>(mean = -4.209, SD = 2.597) | 17 |
| **Proportion with long-COVID** |  |  |  |
| **In adults** | 0.166 | Binomial | 23 |
| **In children (under 20)** | 0.039 | Binomial | 24 |

### Estimation of disease burden

We stratify the disease burden estimates by age group and present it as absolute QALYs lost^25,26^ and QALYs lost per 100,000 persons.

QALYs lost due to premature mortality were calculated as the remaining life expectancy at the time of death per fatal case, i.e., the number of life years lost (LYL). We used nine age groups, in accordance with available Covid-19 mortality statistics: < 10 years, 10-19 years, 20-29 years, 30-39 years, 40-49 years, 50-59 years, 60-69 years, 70-79 years, 80-89 years, and ≥ 90 years, and assumed deaths reported within a given age group occurred at the mid-point of the age interval, in line with previous studies^27,28^.

QALYs lost due to inpatient cases were calculated by multiplying the total number of mild/severe inpatient cases with the disutility per COVID-19 inpatient case. Similarly, QALYs lost due to outpatient and long-COVID cases were calculated by multiplying the total number of outpatient and long-COVID cases with the respective disutilities per case (see table 1).

The total disease burden of COVID-19 was expressed as the sum of the above, i.e., the QALYs lost due to disease in inpatient cases, outpatient cases and due to premature deaths.

Two-sided *p* values of < 0·05 were considered to show statistical significance. All analyses were conducted by R, version 4.1.3^29^.

### Sensitivity analysis

We conducted probabilistic sensitivity analysis to acknowledge uncertainty and examine the robustness of our results. We ran 1,000 simulations of the disease burden estimation with different sets of parameter values derived from the defined ranges and distributions. The range of each parameter, distribution, and the references we used to determine it are available in Table 1. The influence of each parameter on the total disease burden was evaluated by a linear regression analysis with 1,000 simulation results as an independent variable and 1,000 parameter sets as dependent variables.

### Ethics approval

All data used in this study are publicly available. As such, the datasets used in our study were de-identified and fully anonymized. Therefore, this study did not require specific ethical approval.

## Results

In total, 1,728,228 COVID-19 cases were confirmed in Japan from the beginning of 2020 to the end of 2021. Among them, 232,495 cases were observed in 2020 and 1,495,733 cases in 2021. A relatively small number of cases was observed in Waves 1 and 2 (99,959 cases and 1,765 deaths) in Japan, and more than half of the total cases between 2020 and 2021 were observed in Wave 5 (931,393 cases), although the number of deaths in Wave 5 was comparatively small (3,762 deaths). Similarly, the maximum number of cases requiring hospitalization in Wave 1 was 6,250, while in Wave 5 it was 231,596. Figure 2 shows the epidemic curve of confirmed cases and number of cases requiring hospitalization from 2020 to 2021. When the life years lost were adjusted by the age-specific population norm for health-related quality of life (as shown in the Supplementary file), the interpretation of these results did not change.

**Figure 2.**
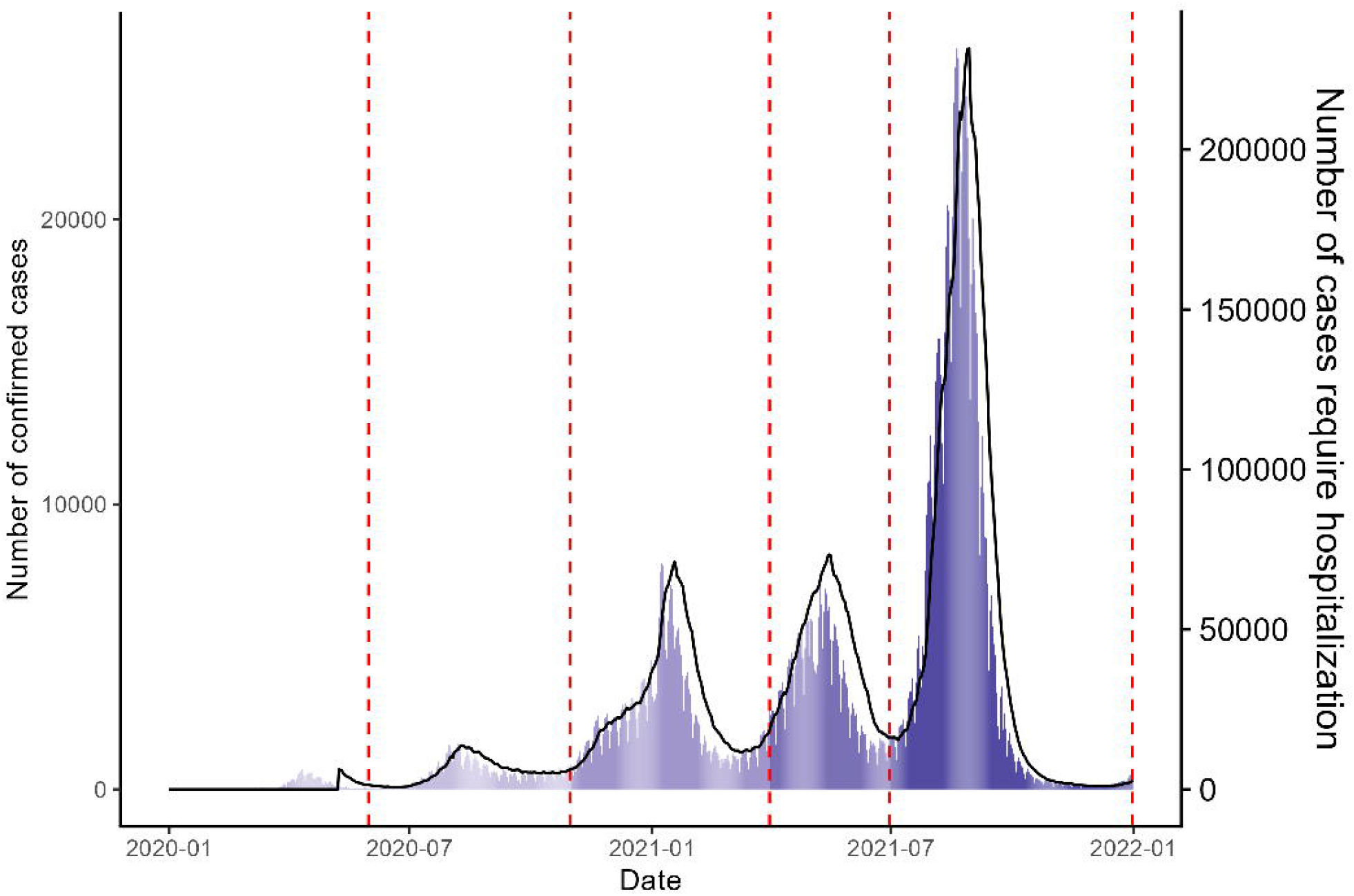
Daily number of confirmed COVID-19 cases and hospitalisations. Wave 1; 01/01/2020-05/31/2020, Wave 2; 06/01/2020-10/31/2020, Wave 3; 11/01/2020-03/31/2021, Wave 4; 4/1/2021-6/30/2021, Wave 5; 7/1/2021-12/31/2021 Bars represent the daily number of confirmed cases. The black line represents the number of cases requiring hospitalization. Vertical lines represent the delimitation of five epidemic waves.

More than a half of total cases were observed in Wave 5 (931,393 cases). The Delta variant of concern (VOC) was dominant in this period^30,31^, as it outcompeted the less transmissible Alpha VOC in July 2021.

The number of fatal cases was 3,095 in 2020 and 14,520 in 2021. The case-fatality ratio (CFR) was 0·013 in 2020 and 0·0097 in 2021, respectively. The estimated average age of fatal cases was 80·2 (Wave 1 and 2), 82·3 (Wave 3), 81·5 (Wave 4), and 75·8 (Wave 5) years.

QALYs lost due to COVID-19 were estimated as 286,781 over two years, or an average of 114·0 QALYs per 100,000 population per year. The observed disease burden differed substantially between waves: from 8·5 QALYs per 100,000 population in Wave 1, up to 96·2 QALYs per 100,000 population in Wave 5. Figure 3 shows the evolution of QALYs lost per 100,000 population in the total population and by broad age group (under 40, 40-69, and over 69) over the waves. The disease burden in younger age groups gradually increased during the study period with 36·4%, 47·6%, 36·6%, 45·0% and 68·2% of the QALYs lost occurring in the age groups younger than 70 in Waves 1 through 5, respectively. When life years lost are adjusted for quality these percentages become38·7%, 50·7%, 39·6%, 48·3%, and 70·7%, respectively.

**Figure 3.**
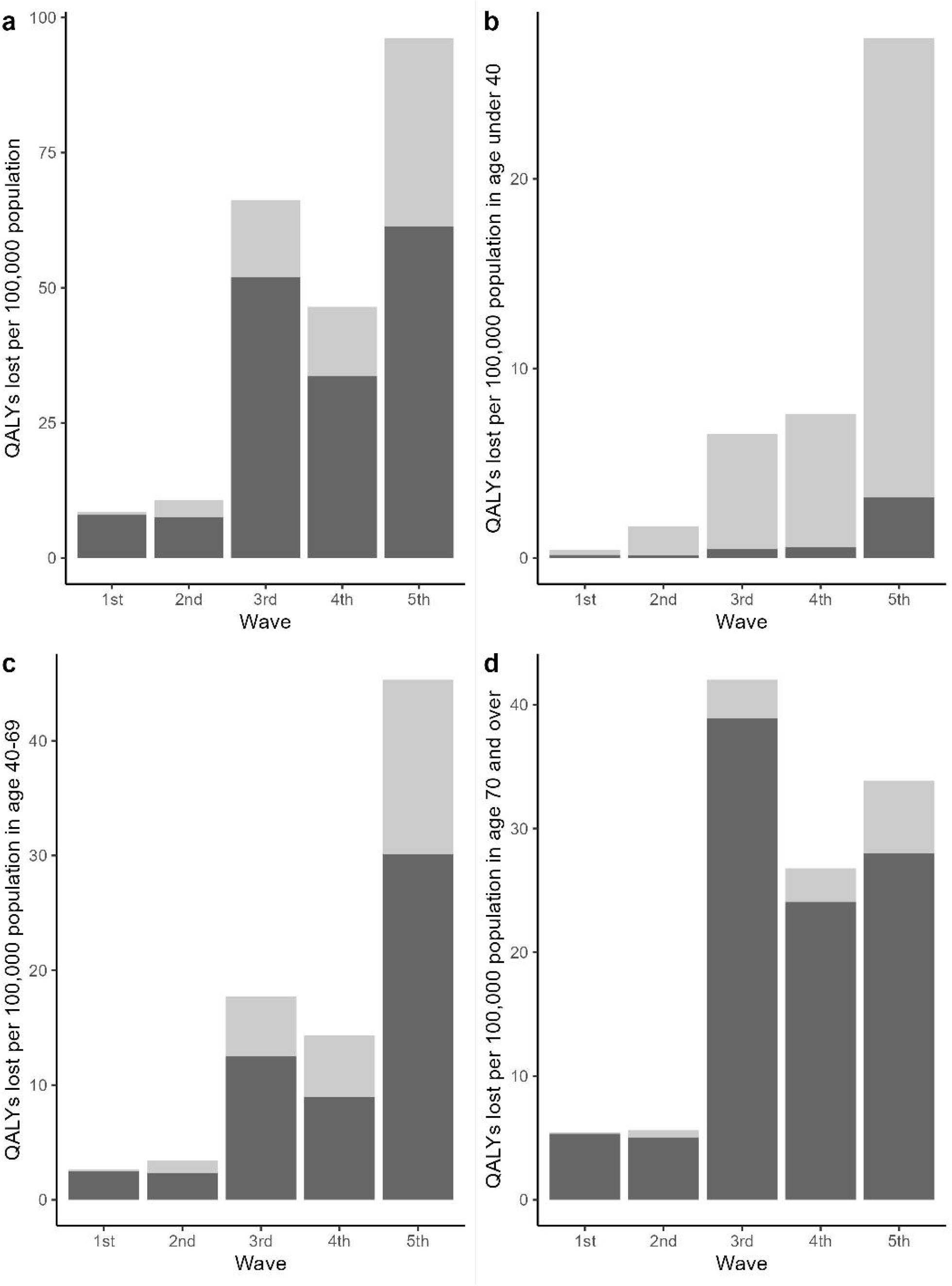
Comparison of QALYs lost per 100,000 population in each epidemic wave. Wave 1; 01/01/2020-05/31/2020, Wave 2; 06/01/2020-10/31/2020, Wave 3; 11/01/2020-03/31/2021, Wave 4; 4/1/2021-6/30/2021, Wave 5; 7/1/2021-12/31/2021 Light grey bars represent QALYs lost due to morbidity and dark grey bars represent QALYs lost due to mortality. QALYs; Quality-adjusted life years Panel a: Disease burden in total population Panel b: Disease burden in population under 40 Panel c: Disease burden in population between age 40 and 69 Panel d: Disease burden in population 70 and over

More than 70% of QALYs were lost due to premature mortality (204,437·2 out of 286,781·7, 71·3%), while nearly 20% were lost due to morbidity in outpatient cases (57031·5 QALYs lost, 19·9%), and only a small part of the burden came from morbidity in severe cases (422·5 QALYs lost, 0·1%). Long-COVID accounts for 3·4% of the total disease burden (9,791·7 QALYs lost). Figure 4 shows the breakdown of disease burden attributed to each clinical status.

**Figure 4.**
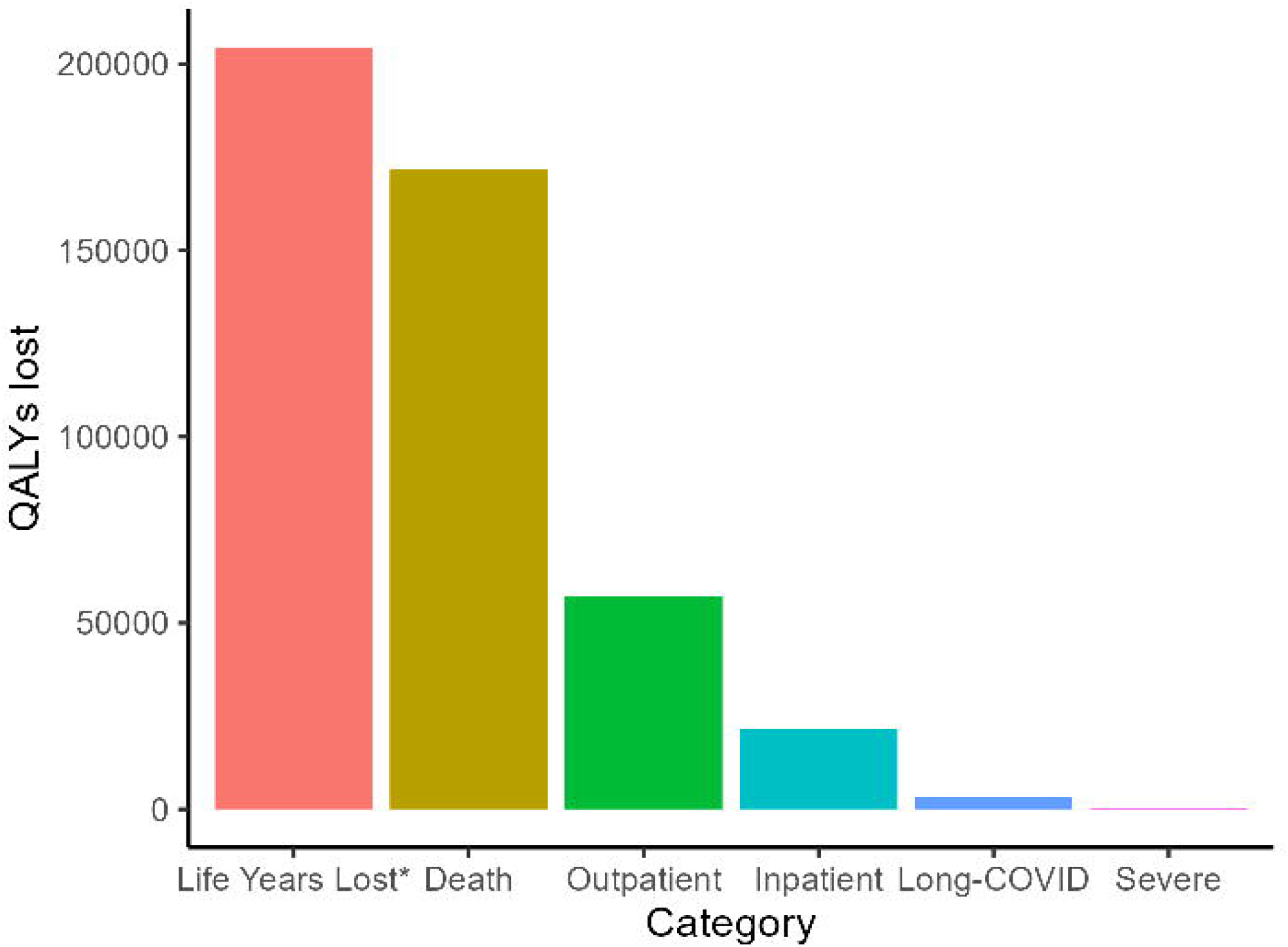
Breakdown of disease burden by clinical manifestation of COVID-19. QALYs; Quality-adjusted life years *Count without quality adjustment (i.e., assuming life years lost due to premature mortality would have been lived in perfect health).

Probabilistic sensitivity analysis showed the disease burden directly due to COVID-19 ranged between 226,883 and 512,236 (median 242,834, IQR 236,108 to 254,488) QALYs, or between 90·1 and 203·6 (median 96·5, IQR 93·8 to 101·1) QALYs per 100,000 population. The disutility per outpatient case was the most influential parameter for the estimated QALY losses due to morbidity, whereas the disutility per long-COVID patient was the second most influential. Figure 5 shows the influence of these and the other input parameters by their coefficient in a linear regression analysis using 1,000 input parameter sets and 1,000 associated QALY estimates. Clearly, accurate estimates for the disutilities per outpatient and per long-COVID patient are important to estimate the burden of disease from COVID-19.

**Figure 5.**
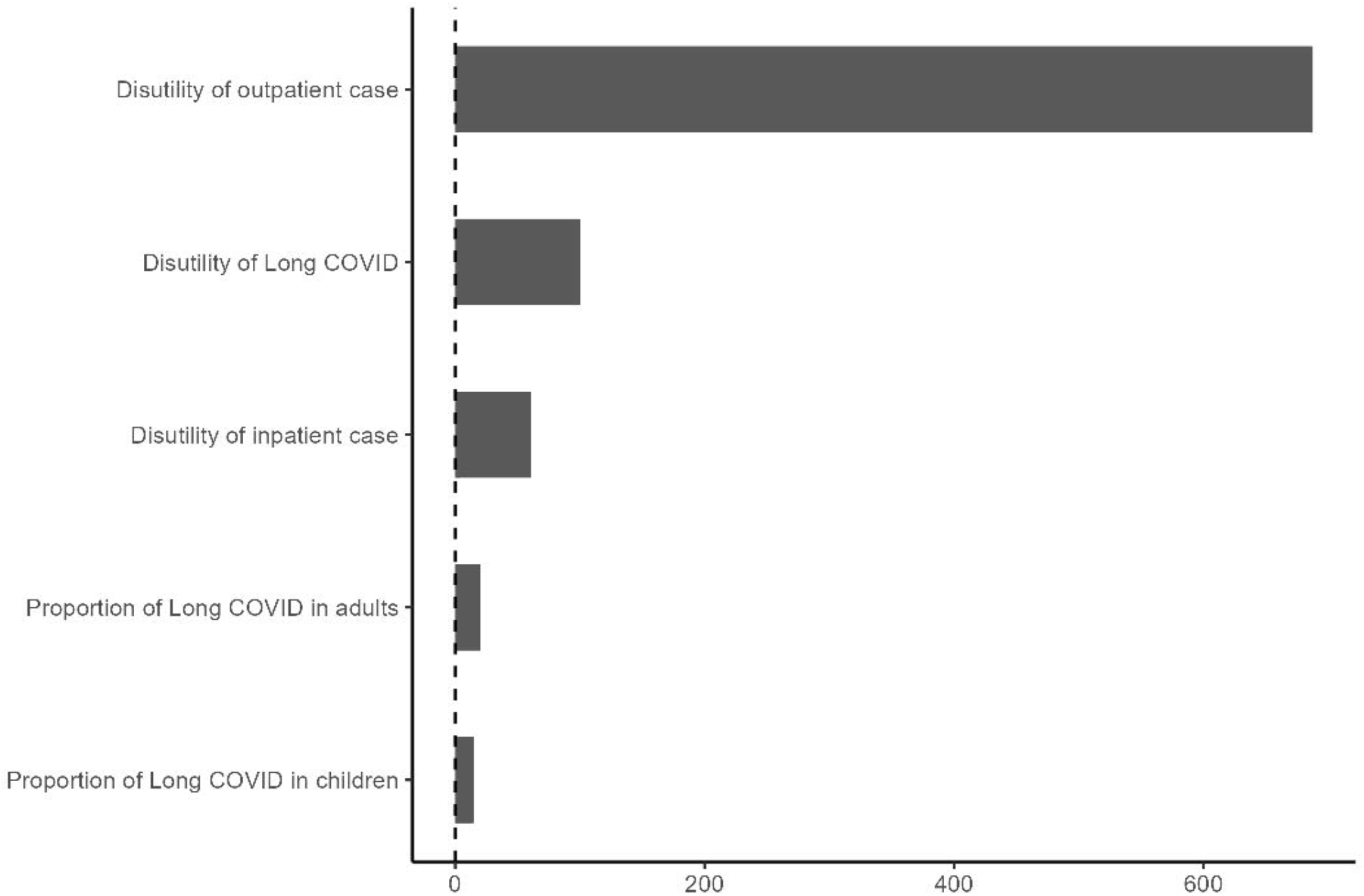
Linear regression coefficient of each parameter included in the probabilistic sensitivity analysis X axis represents coefficient value of each parameter.

## Discussion

To the best of our knowledge, this is the first study which estimated the disease burden directly due to COVID-19 in Japan in the first two years of the pandemic. The disease burden brought by this emerging infectious disease during the first two years was smaller than in most other high-income countries (HICs). For instance, McDonald and colleagues reported that the total disease burden per-capita in the Netherlands in 2020 due to COVID-19 was 1,640 Disability-adjusted life-years (DALYs) per 100,000 population^32^. This number is more than ten times higher than that of our study. Other countries, e.g., Scotland and Malta, also reported similar size of burden in 2020^33,34^. Germany and Denmark reported smaller burden, however, the results were still much larger than ours (368 and 520 DALYs/100,000, respectively)^35,36^.

An obvious difference between previous studies and ours is that we used QALYs instead of DALYs to express burden of disease. Many guidelines advocate the use of a combined measure of morbidity and mortality as preferred outcome in economic evaluation, and the majority of country-specific guidelines, including those for Japan, prescribe the use of QALYs^37^, although some influential generic guidelines such as the IDSI^38^ and the 2019 WHO guide for economic evaluation of vaccinations^39^ indicate that the choice between these outcome measures may depend on the analyst’s preference and of the specific intervention under study. QALYs were used in our study to include the disease burden attributable to long-COVID based on a previous observational Japanese study using the EQ-5D-3L questionnaire^17^. Although this choice may limit the comparability with studies using DALYs as an outcome, it allows us to attain our primary objective, i.e., assessing the disease burden by clinical manifestation, wave and age group, and have a basis to start from to assess the QALY impact of interventions (as one would for economic evaluation). Furthermore, DALYs were conceived in a very similar manner as QALYs, and applied studies using both measures have reported relatively small differences^25^.

As for the total disease burden, this could be simply attributed to the relatively small number of confirmed cases and deaths per population in Japan^11^. Mortality of COVID-19 cases was smaller in Japan than in most other HICs during the study period^19,40–42^. With regard to the number of deaths, the total number of all-cause excess deaths from the beginning of 2020 to the end of 2021 was estimated at between 11,014 and 58,905, while all-cause exiguous deaths in the same period was between 9,069 and 45,185^43^. Considering these numbers included the influence of diseases other than COVID-19, the number of indirect deaths due to COVID-19 might not change the total burden estimates substantially. One can speculate about the reasons why the COVID-19 case and death toll tended to be lower in Japan than in many other HICs. Basically, people who presented fever or any symptoms suspicious of COVID-19 were required to visit one of the designated medical facilities and physicians who diagnosed COVID-19 had to report all COVID-19 cases they diagnosed, implying the potential risk of underestimating the number of cases was low. The cumulative number of COVID-19 cases in Japan increased most steeply after the emergence of the Omicron VOC, and the proportion of the Japanese population with non-vaccine induced immunity against COVID-19 remained low, even in 2021^44^.

Considering the breakdown of QALYs lost, the part of the burden attributable to fatal cases was smaller than that in previous studies. Although only 71·3% of QALYs lost was attributed to fatal cases in our study, more than 90% of the total burden was attributed to LYL in previous studies in other countries^32–36^. It may be partly explained by some studies not including the burden of long-COVID. Furthermore, we have used a simplified approach by counting each lost life year as one, implying that each of these life years was assumed to be lived in perfect health. We also made estimates accounting for a quality adjustment in the life years gained, and as expected, this leads to lower estimates of the QALYs lost due to premature mortality.

The probabilistic sensitivity analysis acknowledges parametric uncertainty and indicates a relatively wide range for the total disease burden. This might be attributed to uncertainty in the estimation of the burden of outpatient cases and long-COVID because we defined the range of disease burden caused by these two statuses according to empirical data in our previous studies^17,21^, then small sample sizes affected their uncertainty. Additionally, it would also be due to the fact that there is no single established definition of long-COVID. Tsuzuki et al defined long-COVID as four weeks or longer duration of symptoms after diagnosis of COVID-19^17^, however, WHO defines post COVID-19 condition as the status “usually 3 months from the onset of COVID-19 with symptoms and that last for at least 2 months and cannot be explained by an alternative diagnosis”^45^. With changing definitions, the proportion of symptomatic cases incurring long-COVID varies significantly. In addition, disutility of each long-COVID case varies substantially by study^46–48^ which will bring further uncertainty.

There are several limitations in this study, similar to the previous studies. First, our results did not consider how many cases were unreported. As described above, all diagnosed COVID-19 cases including asymptomatic ones had to be reported in Japan, nevertheless some level of underreporting seems inevitable. For instance, McKenzie and colleagues insisted that there might be 1·77 times higher number of cases than reported during the same period (from the beginning of 2020 to the end of 2021)^49^. It is difficult for us to estimate the precise cumulative incidence of COVID-19 because we have very little evidence on the seroprevalence of SARS-CoV-2 in Japan^44,49^. However, most of such unreported cases were expected to be “asymptomatic” because they had to be reported if they presented any suspicious symptoms. Since our objective is to assess the disease burden directly attributable due to COVID-19, we can ignore such cases, although they may have given rise to anxiety, especially in 2020, and have an impact on mental wellbeing in both the infected person and their direct contacts in and outside the household.

Second, we did not take the burden to the healthcare systems into consideration. Japanese government decided to admit all the patients diagnosed with COVID-19 regardless of its severity in the early phase of the pandemic^5^. As a result, a large number of tertiary hospitals could not offer normal healthcare due to high burden of the management of COVID-19 cases. This would have brought additional burden to society as previous studies reported^50,51^. Nevertheless, we believe that this additional burden to the healthcare system did not contribute to increase the total disease burden, because excess mortality attributable to the causes other than COVID-19 was smaller than before during the study period^43^, and no large catch-up effect due to postponed care has been detected in mortality statistics to date.

Third, we could not consider the impact of non-pharmaceutical interventions (NPIs). The Japanese government had implemented a “state of emergency” declaration as a kind of recommendation to avoid social contact like capital lockdown in other countries^52^. Such interventions might contribute to decrease the number of COVID-19 cases in exchange for the economic burden. We have scarce evidence about both the economic aspects of these non-pharmaceutical countermeasures so far and a counterfactual scenario about the burden of disease and the economic impact in case we had not implemented such NPIs. Although it seems clear that the total disease burden due to COVID-19 in Japan was relatively smaller than in most other HICs, the impact of the indirect effects of NPIs on the overall disease burden remains a topic on research, as it is in many other countries.

Fourth, we did not distinguish between primary and secondary infections in unvaccinated persons and breakthrough infections in vaccinated persons. The cases reported in 2021, included breakthrough infections, but we lacked the information to specify these cases. Al-Aly et al. showed that the risk of death and post-acute sequelae were higher in unvaccinated cases than breakthrough infections^53^, and the total burden might become slightly smaller if we consider the impact of breakthrough infection. Nevertheless, the number of breakthrough infection can be considered smaller than the normal ones, then its impact also should be a small one.

Fifth, in line with all other studies known to us, we exclude other indirect aspects of the disease burden such as that caused by isolation, imposed on patients’ family members, and so forth, because the number of isolated persons and the number of patients’ household members are not well documented. Although our previous studies had estimated the per-patient impact of these aspects^21,54^, future work will attempt to combine these results with the estimated number of isolated people and household members of infected cases as part of a more general assessment of the indirect burden.

## Conclusions

Most of the disease burden due to COVID-19 in Japan from the beginning of 2020 to the end of 2021 was incurred in 2021 during waves 3, 4, and 5. The proportion of the total burden due to non-fatal disease increased gradually and this was probably due to the lower mortality by COVID-19 in the latter half of the study period, especially in the elderly. It also shows that younger people contributed more to the disease burden as SARS-CoV-2 circulated more in the general population from 2021 onwards. The estimated disease burden in Japan was smaller than in most other HICs and this can be attributed to the small number of confirmed cases in Japan. Future research may further explore the underlying reasons for the lower cumulative incidence of COVID-19 cases in Japan, while taking contextual factors into consideration.

## Supporting information

Supplementary results

## Data Availability

The data that support the findings of this study are freely available through online.

## Acknowledgment

Not Applicable.

## Author contributions

Shinya Tsuzuki: Conceptualization, Funding Acquisition, Data Curation, Formal Analysis, Methodology, Visualization, Original Draft Preparation. Philippe Beutels: Supervision, Project Administration, Review & Editing.

## Data availability

The data that support the findings of this study are freely available through online.

## Ethics approval

This study analysed data which were publicly available. As such, the datasets used in our study were de-identified and fully anonymized in advance, then therefore this this study did not require ethical approval.

## References

1 Guan W, Ni Z, Hu Y, et al. Clinical Characteristics of Coronavirus Disease 2019 in China. N Engl J Med 2020; 382: 1708–20.

2 Huang C, Wang Y, Li X, et al. Clinical features of patients infected with 2019 novel coronavirus in Wuhan, China. The Lancet 2020; 395: 497–506.

3 Chen N, Zhou M, Dong X, et al. Epidemiological and clinical characteristics of 99 cases of 2019 novel coronavirus pneumonia in Wuhan, China: a descriptive study. The Lancet 2020; 395: 507–13.

4 Ministry of Health, Labour and Welfare. Current status of the novel coronavirus infection and the response of the MHLW. https://www.mhlw.go.jp/stf/newpage_09290.html (accessed June 1, 2022).

5 Ministry of Health, Labour and Welfare. Information list for local authorities and healthcare facilities: COVID-19. 2020. https://www.mhlw.go.jp/stf/seisakunitsuite/newpage_00024.html (accessed Aug 17, 2021).

6 Matsunaga N, Hayakawa K, Terada M, et al. Clinical epidemiology of hospitalized patients with COVID-19 in Japan: Report of the COVID-19 REGISTRY JAPAN. Clin Infect Dis 2020; published online Sept. DOI:10.1093/cid/ciaa1470.

7 Inoue H. Japanese strategy to COVID-19: How does it work? Global Health & Medicine 2020; 2: 131–2.

8 Ministry of Health, Labour and Welfare. Information on health and medical consultation. https://www.mhlw.go.jp/stf/covid-19/kenkou-iryousoudan_00006.html (accessed Feb 23, 2022).

9 Yamaguchi S, Okada A, Sunaga S, et al. Impact of COVID-19 pandemic on healthcare service use for non-COVID-19 patients in Japan: retrospective cohort study. BMJ Open 2022; 12: e060390.

10 Ono A, Koizumi R, Tsuzuki S, et al. Antimicrobial Use Fell Substantially in Japan in 2020—The COVID-19 Pandemic May Have Played a Role. International Journal of Infectious Diseases 2022. DOI:10.1016/j.ijid.2022.03.019.

11 Coronavirus (COVID-19) Cases - Our World in Data. https://ourworldindata.org/covid-cases (accessed Feb 24, 2022).

12 Healthcare in the UK | Coronavirus in the UK. https://coronavirus.data.gov.uk/details/healthcare (accessed Dec 3, 2022).

13 United States - COVID-19 Overview - Johns Hopkins. Johns Hopkins Coronavirus Resource Center. https://coronavirus.jhu.edu/region/united-states (accessed Dec 3, 2022).

14 Idogawa M, Tange S, Nakase H, Tokino T. Interactive Web-based Graphs of Coronavirus Disease 2019 Cases and Deaths per Population by Country. Clinical Infectious Diseases 2020; 71: 902–3.

15 del Rio C, Collins LF, Malani P. Long-term Health Consequences of COVID-19. JAMA 2020; published online Oct 5. DOI:10.1001/jama.2020.19719.

16 Hayakawa K, Asai Y, Matsunaga N, et al. Evaluation of the representativeness of data in the COVID-19 Registry Japan during the first six waves of the epidemic. Global Health & Medicine 2022. DOI:10.35772/ghm.2022.01033.

17 Tsuzuki S, Miyazato Y, Terada M, Morioka S, Ohmagari N, Beutels P. Impact of long-COVID on health-related quality of life in Japanese COVID-19 patients. Health and Quality of Life Outcomes 2022; 20: 125.

18 Miyazato Y, Morioka S, Tsuzuki S, et al. Prolonged and Late-Onset Symptoms of Coronavirus Disease 2019. Open Forum Infect Dis 2020; 7: ofaa507.

19 Ministry of Health, Labour and Welfare. Visualizing the data: information on COVID-19 infections. https://covid19.mhlw.go.jp/extensions/public/en/index.html (accessed Feb 24, 2022).

20 National Institute of Population and Social Security Research. Population Statistics. 2020. http://www.ipss.go.jp/syoushika/tohkei/Popular/Popular2014.asp?chap=5&title1=%87X%81D%8E%80%96S%81E%8E%F5%96%BD (accessed Dec 20, 2020).

21 Tsuzuki S, Ohmagari N, Beutels P. The burden of isolation to the individual: a comparison between isolation for COVID-19 and for other influenza-like illnesses in Japan. Epidemiology and Infection 2022; 150: e5-undefined.

22 Glick HA, Miyazaki T, Hirano K, et al. One-Year Quality of Life Post–Pneumonia Diagnosis in Japanese Adults. Clinical Infectious Diseases 2021; 73: 283–90.

23 Thompson EJ, Williams DM, Walker AJ, et al. Long COVID burden and risk factors in 10 UK longitudinal studies and electronic health records. Nature Communications 2022; 13. DOI:10.1038/s41467-022-30836-0.

24 Wulf Hanson S, Abbafati C, Aerts JG, et al. Estimated Global Proportions of Individuals With Persistent Fatigue, Cognitive, and Respiratory Symptom Clusters Following Symptomatic COVID-19 in 2020 and 2021. JAMA 2022. DOI:10.1001/jama.2022.18931.

25 Augustovski F, Colantonio LD, Galante J, et al. Measuring the Benefits of Healthcare: DALYs and QALYs - Does the Choice of Measure Matter? A Case Study of Two Preventive Interventions. Int J Health Policy Manag 2017; 7: 120–36.

26 Drummond MF, Sculpher MJ, Claxton K, Stoddart GL, Torrance GW. Methods for the Economic Evaluation of Health Care Programmes, 4th edn. Oxford: Oxford University Press, 2015.

27 Tsuzuki S, Baguelin M, Pebody R, van Leeuwen E. Modelling the optimal target age group for seasonal influenza vaccination in Japan. Vaccine 2020; 38: 752–62.

28 Tsuzuki S, Schwehm M, Eichner M. Simulation studies to assess the long-term effects of Japan’s change from trivalent to quadrivalent influenza vaccination. Vaccine 2018; 36: 624–30.

29 R Core Team. R: A Language and Environment for Statistical Computing. 2018.

30 Shoji K, Akiyama T, Tsuzuki S, et al. Clinical characteristics of COVID-19 in hospitalized children during the Omicron variant predominant period. J Infect Chemother 2022; 28: 1531–5.

31 Shoji K, Akiyama T, Tsuzuki S, et al. Comparison of the clinical characteristics and outcomes of COVID-19 in children before and after the emergence of Delta variant of concern in Japan. J Infect Chemother 2022; 28: 591–4.

32 Mcdonald SA, Lagerweij GR, De Boer P, et al. The estimated disease burden of acute COVID-19 in the Netherlands in 2020, in disability-adjusted life-years. European Journal of Epidemiology 2022. DOI:10.1007/s10654-022-00895-0.

33 Wyper GMA, Fletcher E, Grant I, et al. Measuring disability-adjusted life years (DALYs) due to COVID-19 in Scotland, 2020. Archives of Public Health 2022; 80. DOI:10.1186/s13690-022-00862-x.

34 Cuschieri S, Calleja N, Devleesschauwer B, Wyper GMA. Estimating the direct Covid-19 disability-adjusted life years impact on the Malta population for the first full year. BMC Public Health 2021; 21. DOI:10.1186/s12889-021-11893-4.

35 Rommel A, von der Lippe E, Pla⍰ D, et al. The COVID-19 Disease Burden in Germany in 2020. Dtsch Arztebl International 2021; 118: 145–51.

36 Pires SM, Redondo HG, Espenhain L, et al. Disability adjusted life years associated with COVID-19 in Denmark in the first year of the pandemic. BMC Public Health 2022; 22. DOI:10.1186/s12889-022-13694-9.

37 Zhao Y, Feng H, Qu J, Luo X, Ma W, Tian J. A systematic review of pharmacoeconomic guidelines. Journal of Medical Economics 2018; 21: 85–96.

38 Health Technology Assessment Toolkit. iDSI. 2017; published online Sept 7. https://www.idsihealth.org/htatoolkit/ (accessed Dec 8, 2022).

39 World Health Organization. WHO guide for standardization of economic evaluations of immunization programmes, 2nd ed. https://www.who.int/publications-detail-redirect/who-guide-for-standardization-of-economic-evaluations-of-immunization-programmes-2nd-ed (accessed Dec 8, 2022).

40 Eurosurveillance | Estimates of mortality attributable to COVID-19: a statistical model for monitoring COVID-19 and seasonal influenza, Denmark, spring 2020. https://www.eurosurveillance.org/content/10.2807/1560-7917.ES.2021.26.8.2001646 (accessed Feb 26, 2021).

41 Lipsitch M. Estimating case fatality rates of COVID-19. The Lancet Infectious Diseases 2020; 20: 775.

42 Pastor-Barriuso R, Pérez-Gómez B, Hernán MA, et al. Infection fatality risk for SARS-CoV-2 in community dwelling population of Spain: nationwide seroepidemiological study. BMJ 2020; 371: m4509.

43 Kawashima T, Nomura S, Tanoue Y, et al. Excess All-Cause Deaths during Coronavirus Disease Pandemic, Japan, January–May 2020 - Volume 27, Number 3— March 2021 - Emerging Infectious Diseases journal - CDC. DOI:10.3201/eid2703.203925.

44 Sugiyama A, Okada F, Abe K, et al. A longitudinal study of anti-SARS-CoV-2 antibody seroprevalence in a random sample of the general population in Hiroshima in 2020. Environmental Health and Preventive Medicine 2022; 27: 30–30.

45 World Health Organization. A clinical case definition of post COVID-19 condition by a Delphi consensus, 6 October 2021. 2021; published online Oct 6. https://www.who.int/publications-detail-redirect/WHO-2019-nCoV-Post_COVID-19_condition-Clinical_case_definition-2021.1 (accessed Sept 19, 2022).

46 Sandmann FG, Tessier E, Lacy J, et al. Long-Term Health-Related Quality of Life in Non-Hospitalized Coronavirus Disease 2019 (COVID-19) Cases With Confirmed Severe Acute Respiratory Syndrome Coronavirus 2 (SARS-CoV-2) Infection in England: Longitudinal Analysis and Cross-Sectional Comparison W. Clinical Infectious Diseases 2022. DOI:10.1093/cid/ciac151.

47 Malik P, Patel K, Pinto C, et al. Post-acute COVID-19 syndrome (PCS) and health-related quality of life (HRQoL)-A systematic review and meta-analysis. J Med Virol 2022; 94: 253–62.

48 Malinowska A, Muchlado M, Ślizień Z, et al. Post-COVID-19 Sydrome and Decrease in Health-Related Quality of Life in Kidney Transplant Recipients after SARS-COV-2 Infection-A Cohort Longitudinal Study from the North of Poland. J Clin Med 2021; 10: 5205.

49 Mckenzie L, Shoukat A, Wong KO, et al. Inferring the true number of SARS-CoV-2 infections in Japan. Journal of Infection and Chemotherapy 2022. DOI:10.1016/j.jiac.2022.08.002.

50 Verelst F, Kuylen E, Beutels P. Indications for healthcare surge capacity in European countries facing an exponential increase in coronavirus disease (COVID-19) cases, March 2020. Eurosurveillance 2020; 25: 2000323.

51 McCabe R, Kont MD, Schmit N, et al. Modelling intensive care unit capacity under different epidemiological scenarios of the COVID-19 pandemic in three Western European countries. International Journal of Epidemiology 2021; published online April. DOI:10.1093/ije/dyab034.

52 Machida M, Wada K. Public health responses to COVID-19 in Japan. Global Health & Medicine 2022; 4: 78–82.

53 Al-Aly Z, Bowe B, Xie Y. Long COVID after breakthrough SARS-CoV-2 infection. Nat Med 2022; 28: 1461–7.

54 Tsuzuki S, Yoshihara K. The characteristics of influenza-like illness management in Japan. BMC Public Health 2020; 20: 568.

