## Supplementary results for "The estimated disease burden of COVID-19 in Japan from 2020 to 2021"

**The supplementary file of**

**Table S1. Additional results with adjusted Life Years Lost by the age-specific population norm for quality of life**^1,2^

|  | **QALYs lost** | | | |
| --- | --- | --- | --- | --- |
| **Two years total** | **260,426·2** | | | |
| **Per 100,000 population per year** | **103·5** | | | |
| **Each clinical status:**  **Fatal cases**  **Outpatient cases**  **Severe cases** | **171,804·8**  **57,031·5**  **422·5** | | | |
|  | **Age group** | | | |
| **Each epidemic wave:**  **Wave 1**  **Wave 2**  **Wave 3**  **Wave 4**  **Wave 5** | **Under 40**  **529·7**  **2083·8**  **8173·0**  **9511·1**  **34176·3** | **40-69**  **2995·1**  **3999·0**  **20423·7**  **16675·6**  **52716·9** | **70 and over**  **5591·9**  **5914·2**  **43646·3**  **27995·2**  **35991·4** | **Total**  **9109·0**  **11958·9**  **72207·6**  **51398·1**  **109784·5** |

1	Briggs AH, Goldstein DA, Kirwin E, *et al·* Estimating (quality‐adjusted) life‐year losses associated with deaths: With application to COVID‐19· *Health Economics* 2021; **30**: 699–707.

2	Shiroiwa T, Noto S, Fukuda T· Japanese Population Norms of EQ-5D-5L and Health Utilities Index Mark 3: Disutility Catalog by Disease and Symptom in Community Settings· *Value in Health* 2021; **24**: 1193–202.

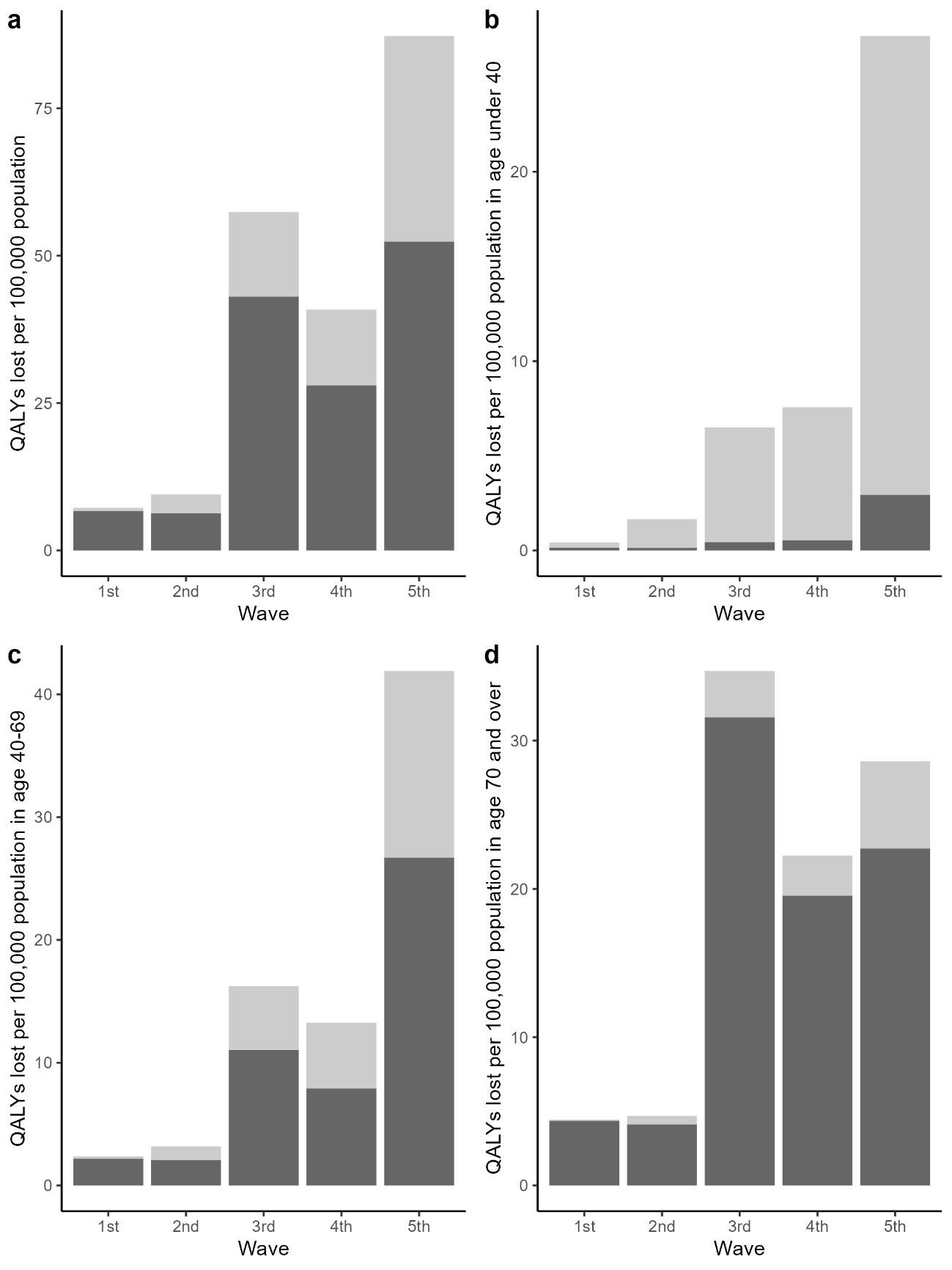

**Figure S1. Wave-specific disease burden with Life Years Lost adjusted by the age-specific population norm for quality of life**

Light grey bars represent QALYs lost due to morbidity and dark grey bars represent QALYs lost due to mortality.
